# An Investigation of Lead Concentration in the Breathing Air and the Blood of Automobile Welders in Birjand, Iran

**DOI:** 10.1101/2022.08.21.22279034

**Authors:** Omolbanin Motamedrezaei, Abdolah Gholami, Gholamreza Sharifzadeh, Farnaz Jahani, Hamed Lotfi

## Abstract

**Introduction:** Lead is known as one of the most dangerous toxic metals in the world and its entry into the body can cause acute and chronic intoxication with a wide range of systemic symptoms. Our goal in the current research is to study the level of lead in the breathing zone and blood of the automobile welders in Birjand.

**Methods:** A cross-sectional, descriptive-analytic study was conducted on 47 automobile welders. The criteria for entering the study included, male gender, having at least 1 year of experience in automobile welding, and at least 8 hours of daily employment in welding. The general information required was collected through a questionnaire. At the beginning of the study, in each working environment air samples were taken in accordance with NIOSH 7082 standards. The analysis of the air lead concentration (ALC) was carried out by a flame atomic absorption spectrophotometer. The blood lead concentration (BLC) was measured by NIOSH 8003 method via graphite furnace atomic absorption spectrophotometer.

**Results:** Based on the data, 12.8% of the workers were smokers and 66% used appropriate personal protective equipment. The average ALC was 0.0458 ± 0.0296 mg/m^3^ and the average BLC of automobile welders was 9.89 ± 7.32 μg/dL. Although Pearson correlation coefficient showed a positive correlation between ALCs and BLCs, this correlation was not statistically significant (p = 0.38, r = 0.18).

**Conclusion:** The findings of this study showed that the average BLC in cigarette smokers and those who did not use PPE was higher than other people; besides, it was higher in individuals over the age of 30 than those under this age. The mean BLC in automobile welders and the mean ALC corresponded to the standards of the American Conference of Governmental Industrial Hygienists (ACGIH).

## Introduction

Lead (Pb) is an ubiquitous toxic metal with several useful properties i.e. high density, ductility, high malleability, resistance to corrosion, low melting point, and recyclability. Lead is extensively used in industries and household appliances, including automobiles, soldering, welding, lead-acid batteries, plastics, paints, fuel, ceramics, food storage cans, etc ^1,2^. Lead has a non-biodegradable nature and remain persistent in the environment ^3^. Although the poisoning features of lead have been detected as early as 370 BC, it still remains an important growing occupational and environmental health concern ^4^. Lead enters the body mainly through inhalation, ingestion, and skin. Inorganic lead has a wide range of adverse effects in humans even at low blood lead concentration (BLC), and there is no safety threshold for that ^5^. Respiratory and gastrointestinal tracts are the main routs of inorganic lead absorption, so workers’ exposure to lead mainly occur through the respiratory tract (40% of inhaled lead is absorbed into the blood plasma). The circulating lead is bound to erythrocytes for about 30 days, and is therefore distributed in several organs, such as the kidneys, liver, brain, bones, teeth, etc ^6^. After exposure, the half-life of lead elimination from blood would be about 30 days, but it remains in bones for 10-20 years; So, BLC could be considered as an indicator for recent lead exposure ^7^.

Welding is the process of bonding two or more metals by applying heat. About 11 million people work as welders in the world; besides, 110 million people are continuously exposed to various levels of welding fumes released during welding process at work. The welding fumes are aerosols consisting of different metals such as lead, ferrous, copper, nickel, magnesium, zinc, cobalt, cadmium, titanium, etc ^8,9^. Lead intoxication could be either acute or chronic. Chronic exposure to lead might have deleterious effects on nervous, renal, hematopoietic, gastrointestinal, reproductive, cardiovascular, immune systems, and also might cause behavioral dysfunctions ^10^. Generally, these adverse effects of lead in the body is associated with systemic inflammation, cellular oxidative stress, and suppressing the antioxidant enzymes, resulting in impairment of cellular functions and/or signaling cascades and cell death ^11^. The clinical manifestations of toxicity caused by lead, include dizziness, anxiety, dementia, muscle weakness, anemia, abdominal colic, constipation, loss of appetite, myalgia, encephalopathy, seizure, and even coma. Additionally, characteristic features are blue lines named Burton’s line, on the gums, wrist drop, and basophilic stippling, which are not necessarily observed in every case of lead intoxication ^12^, moreover, there is limited evidence indicating that among the exposed workers, inorganic lead compounds have carcinogenic effects on the lungs and the stomach ^1,13^. It should be noted that the chance of elevated BLC in children whose fathers have been exposed to lead in workplaces has been higher than those whose fathers been employees ^14^.

Although comprehensive researches have been conducted in the welding industry in IRAN, there has been no study done on the welders in Birjand. As the welders are continuously exposed to the fumes, in the current study we have attempted to investigate the breathing air lead concentrations (ALCs) and BLCs of the automobile welders in Birjand; hence, the results can provide the information needed for assessing the risk of occupational health hazards.

## Materials and methods

### Subject

In this cross-sectional, descriptive-analytic study, 47 automobile welders in Birjand city were studied in the time span of September 2018 to March 2019; the main objective was to estimate ALCs and BLCs of automobile welders in this city (the capital of southern Khorasan province in Iran). Using the Cochran formula, a total sample size of 47 welders was calculated with an error level of 5%. Inclusion criteria consisted of male automobile welders who had at least 1 year of welding experience, and at least 8 hours of daily exposure. We also collected environmental data, including appropriate ventilation of workplace and presence/absence of personal protective equipment (PPE), and demographic data including age, work experience, cigarette smoking, and using PPE with the maximum privacy.

### Personal sampling and measurement of ALC

Air sampling was performed from one site of each two welders’ sites. Personal air sampling was performed to assess the rate of exposure. The occupational exposure limit of lead in the breathing zone is 0.05mg/m^3^, according to standards of the American Conference of Governmental Industrial Hygienists (ACGIH).^15^

The National Institute for Occupational Safety and Health (NIOSH) has developed methods for measuring airborne lead. In this study, the method number 7082 of NIOSH was used for air sampling.^16^ According to this method, first of all a Mixed Cellulose Ester filter (37mm, 0.9 μm pore size) was placed inside the filter holder, and was connected by flexible pipes to a personal air sampling pump (224-44MTX, SKC, US) with a flow rate of 2 L/minute, calibrated by digital calibrator (200-510M, UK), and the filter holder was attached to the collar of the welders (breathing zone). It is noteworthy that to cover the entire work shift of each welder, 3 samples were used at intervals of 2.5 hours. At the end of the shift, the samples were transferred to the laboratory and were analyzed using flame atomic absorption spectrophotometer (Varian AA240, Australia).^17^ Later, 76 samples were selected as controls to eliminate possible error rate during sampling or transferring. The control samples were opened and closed in the sampling environment, and then were transferred to the laboratory like the study samples. Due to the difference between temperature and pressure of the working environment with standard conditions (25°C temperature, and 760 mm Hg pressure), temperature and humidity corrections were done on the sampled air, and then the obtained results were compared with the occupational exposure limit of lead.^18^

### Measurement of BLC

8cc of the peripheral blood samples, collected in vacuum plastic tubes, containing heparin, were transferred to the laboratory immediately and were kept at −80°C until the testing time. BLCs, were measured using graphite furnace atomic absorption spectrophotometer (4-alpha, UK) with a wave-length of 283.3 nm, according to the Method Number 8003 of NIOSH.^19^ According to standards of the ACGIH, the occupational exposure limit of blood lead was 20 μg/dL.^15^

ALCs and BLCs were measured at the Research Laboratory of Birjand University of Medical Sciences. Finally, the obtained results of the blood and air samples were compared with the standards of the ACGIH; and the association between demographic variables and BLCs was investigated.

### Ethical considerations and informed consent

The protocol of study was approved by the Committee on Research Ethics at Birjand University of Medical Sciences with the approved No. 4794 (ethics code IR.Bums.REC.1396.360). Before entering the study, welders completed an informed consent form; besides, they were assured that all welders’ information would remain confidential with the research team.

### Statistical analysis

Data analysis was done using SPSS software Version 18, and due to the normal distribution of lead levels, independent T-test was used at α level of 0.05. Descriptive statistical tests were performed for quantitative variables, mean, and standard deviation either. The data were shown as Mean ± SD. The significance threshold was considered less than 0.05.

## Results

### Environmental and personal demographic information

The data on 47 welders in Birjand city were collected in the time span of September 2018 to March 2019. The welders’ mean age was 32.6 ± 8.8 years (range: 17 to 39 years), and the mean welding experience was 11.1 ± 7.6 years (range: 1 to 40 years); 12.8% of them were cigarette smokers, and 66% used PPE (Table 1). Because of the sameness of ventilation condition, ventilation cannot be considered as one of the factors of evaluation of BLC; as there was no local ventilation in any workplace, and the general ventilation was the same and was done by fan and natural ventilation.

**Table 1.** Environmental and individual demographic information of the welders.

| Variable | Frequency | Percentage |
| --- | --- | --- |
| Age (years) |  |  |
| Minimum | 17 | - |
| Maximum | 39 | - |
| Mean | 32.6 ± 8.8 | - |
| Welding experience (years) |  |  |
| Minimum | 1 | - |
| Maximum | 40 | - |
| Mean | 11.1 ± 7.6 | - |
| Ventilation |  |  |
| Appropriate | 47 | 100 |
| Inappropriate | 0 | 0 |
| Cigarette smoking |  |  |
| Yes | 6 | 12.8 |
| No | 41 | 87.2 |
| Using PPE |  |  |
| Yes | 31 | 66 |
| No | 16 | 34 |

### Lead concentration in the air and blood samples

Air samples were taken at 26 sites of welders’ workplace. The maximum ALC was 0.115 mg/m^3^, and the mean was 0.0458 ± 0.0296 mg/m^3^. The maximum BLC was 28.89 μg/dL, and the mean was 9.89 ± 7.32 μg/dL (Table 2). Pearson correlation coefficient (r) showed a positive correlation between ALCs and BLCs; However, this correlation was not statistically significant (P= 0.38, r= 0.18).

**Table 2.** Lead concentration in the air and welders’ blood samples.

| Variable | Frequency | Minimum | Maximum | Mean | Median | Standard Deviation |
| --- | --- | --- | --- | --- | --- | --- |
| Air lead concentration (mg/m<sup>3</sup>) | 26 | 0 | 0.115 | 0.0458 | 0.041 | 0.0296 |
| Blood lead concentration (µg/dL) | 47 | 0 | 28.89 | 9.89 | 9.33 | 7.32 |

### Relationship between demographic variables and BLC

Table 3 shows the relationship between demographic variables and the BLCs. Analysis on the bias of using PPE, smoking cigarette, and age showed no significant difference among the welders; however, the mean BLC was higher in cigarette smokers and those who did not use PPE. Additionally, the mean BLC in welders older than 30 years was higher than the mean BLC in welders younger than 30.

**Table 3.** Comparison of the mean BLCs of welders in terms of demographic variables.

| Variable | Frequency | Mean ± SD | Independent T-test |
| --- | --- | --- | --- |
| Using PPE |  |  |  |
| Yes | 31 | 0.0968 ± 0.075 | t= 0.28 |
| No | 16 | 0.1031 ± 0.071 | P= 0.78 |
| Cigarette smoking |  |  |  |
| Yes | 6 | 0.1282 ± 0.082 | t= 1.05 |
| No | 41 | 0.0946 ± 0.072 | P= 0.3 |
| Age |  |  |  |
| ≤30 years | 22 | 0.0907 ± 0.078 | t= 0.72 |
| >30 years | 25 | 0.1061 ± 0.069 | P= 48 |

### Comparison of ALCs and BLCs with the standards of the ACGIH

According to the data, shown in Table 4, the ALCs were complied with the standards of the ACGIH; however, the BLCs of welders were significantly lower than the standards of the ACGIH (P< 0.001).

**Table 4.** Comparison of ALCs and BLCs with the standards of the ACGIH.

| Variable | Frequency | Mean ± SD | One sample T-test |
| --- | --- | --- | --- |
| Air lead concentration<br>(mg/m<sup>3</sup>) | 26 | 0.0458 ± 0.0296 | t= 0.72<br>P= 0.48 |
| Blood lead concentration<br>(μg/dL) | 47 | 9.89 ± 7.32 | t= 6.46<br>P< 0.001 |

## Discussion

Occupational exposure to toxic metals is a global concern for workers employed in polluting industries, in the industrial hygiene field ^20^. The fields around the welders (breathing zone) contain the contaminants produced during welding operations ^21^. Lead is one of these pollutants, and 40% of inhaled lead is absorbed to blood circulation, which is the main rout of inorganic lead absorption ^6,22^. in the present study, we have investigated the ALCs and BLCs of automobile welders in Birjand city. Base on the results of this research, the mean BLC of automobile welders and the mean ALC were significantly lower than the permissible limit recommended by ACGIH; moreover, Pearson correlation coefficient demonstrated a positive correlation (r = 0.18) between ALCs and BLCs. A significant correlation between ALC and BLC of the workers being occupationally exposed to lead-containing aerosols, has been reported by Ono (2021) and Pierre (2002) ^23,24^.

In this study air sampling was performed at 26 sites. The maximum ALC was 0.115 mg/m^3^, and the mean ALC was 0.0458 ± 0.0296 mg/m^3^, which base on the standards of ACGIH were lower than the permissible limit (0.05 mg/m^3^). To investigate the influence of being exposed to airborne lead on BLCs, a cross-sectional study done by Odongo et al.^25^, (2020) among twenty automobile repair artisans in Kenya. Their findings showed that the mean ALC of workers was 22.55 ± 5.05 μg/m^3^, which was lower than the permissible limit; therefore, this study is consistent with the results of our study. On the other hand, Ithnin et al.^26^, (2019) conducted a cross-sectional study to assess the effects of being exposed to welding fumes on the lungs functionality tests of 30 welders in Malaysia. It was shown that the mean ALC of workers’ breathing zone was 2.752 mg/m^3^, which was higher than the permissible limit. The results of this study are inconsistent with the present study. This disparity might be due to the type of equipments used, as well as the different ventilation conditions.

In the present study, the maximum BLC was 28.89 μg/dL, and the mean BLC of 47 welders was 9.89 ± 7.32 μg/dL; according to the standards of the ACGIH, the mean BLC was significantly lower than the permissible limit (20 μg/dL). In line with our study, Goyal et al.^27^, (2021) conducted a study with the aim of measuring the blood concentration of lead and cadmium in 207 individuals in Jodhpur, India. It was shown that the mean BLC of welders was 7.97 ± 1.92 μg/dL, while in non-exposed population the mean BLC was 1.09 ± .073 μg/dL, this difference was statistically significant, but the reported BLC was lower than the permissible limit recommended by ACGIH.^15^ Another cross-sectional study done by Ono et al.^23^, (2014) to investigate the association between ALC and BLC in workers of lead-acid battery factory in JAPAN, which showed that the mean BLC was 10.2 μg/dL, which was lower than the permissible limit. Similar findings were shown by Shriadeh et al.^28^, (2018); compared to controls, they indicated a significantly higher BLC of automobile workers in Jordan. Based on their results, BLC of automobile welders was 14.5 ± 1.4 μg/dL, which was lower than the permissible limit. Another study done in other province reported similar results either. Kalantari et al.^29^, (2009) conducted a case control study to evaluate the BLC and the risk of lead intoxication for 80 workers of zinc melting factory of Dandi in Zanjan. It was shown that the mean BLC of the workers was 16.06 μg/dL, which was lower than the permissible limit. The results of these studies is consistent with the results of the current study. On the other hand, a cross-sectional study was done by Dehghan et al.^9^, (2019) to investigate the association between BLC and ALC with reproductive hormones of 85 welders working in a water transfer company in Iran. It was found that the mean BLC among welders was 460.28 ± 93.65 μg/L which was significantly higher than the permissible limit. Additionally, a cross-sectional, descriptive-analytic study was done by Mirsalimi et al.^30^, (2019) to measure the BLC of 46 workers employed in the lead and zinc mine in Isfahan province. According to their results, the mean BLC of workers was 24.5 μg/dL, which was higher than the permissible limit. In those studies, the BLCs were higher than the welders described in the present study; this might be due to the higher air pollution in Tehran and Isfahan as populous industrial cities ^31,32^. Another cross-sectional study done by Ahmad et al.^33^, (2014) to investigate the BLC, and health problems related to lead intoxication in workers of lead-acid battery factory in Bangladesh, which showed that the BLCs were high among workers, and the mean BLC was 65.25 ± 26.66 μg/dL. The results of the mentioned studies is not consistent with our study. This is probably due to the exposure of welders to other sources of lead; furthermore, it should be considered that other causes such as being exposed to leaded paints, coal combustion, and other lead-containing chemical derivates, how to use PPE, opium addiction, air pollution, using traditional medicine, and even type of diet could elevate the BLC ^34^. Additionally, hygienic principles mostly are not hold by some workers; thus the risk of over-exposure to lead is increased. These workers often don’t use mask, goggles, gloves, and aprons. Many of them eat food without washing their hands before, and also eat and smoke in their workplaces, so this condition and carelessness in holding hygienic principles would expose them to lead through ingestion, inhalation and skin contact ^35^.

Elevated BLCs in some of the welders participating in our study may be related to grounded water and some food contamination with lead in Birjand ^36^. In this regard, Mansouri et al.^37^, (2012) showed that the concentration of lead in groundwater of the Birjand flood plain was 0.023 mg/L, which was higher than the national and international guidelines. Besides, a study conducted by Zeinali, et al.^38^, (2019) to evaluate heavy metals concentrations in the meat and other edible organs of cow and sheep in Birjand. It was found that all samples were contaminated with lead, as well as other heavy metals. Moreover, it has been reported in another study that all edible organs of chicken in Birjand were contaminated with lead ^39^.

It is widely believed that tobacco products contain lead, and this metal can be easily transferred to the body via cigarettes smoking, so smokers have higher BLCs than non-smokers ^40,41^. in the present study, the mean BLC of cigarette smokers was higher than other welders, but this difference was not statistically significant. Shakeri, et al.^42^, (2018) conducted a study to compare levels of different heavy metals between smokers and non-smokers in Birjand city. It was shown that there was no significant difference between the mean BLC of two groups. In line with our study, Ono (2021) found no significant difference between BLC in smoker and non-smoker workers in Japan ^23^; moreover, a study already done in Tunisia (2019) on battery manufacturing workers showed that there was no significant difference between the BLC of cigarette-smoker and non-smoker workers. The close association between cigarette smoking habit and BLC in some studies could be attributed to some confounding factors ^43^. The effects of smoking habit on BLC need to be more considered in future studies.

In fact, older people have been exposed to lead for longer time, so they are expected to have higher BLCs ^44^. In the present study the BLC of welders older than 30 years old was higher than the BLC of welders under 30 years old, though it was not statistically significant. A study was carried out by Singh et al.^45^, (2021) to evaluate the levels of heavy metals in occupationally exposed workers in India. It was reported higher BLCs in older workers. Similar findings were observed in another study; according to the report of which, among battery factory workers, with increase in age the BLCs were higher either ^46^. Mansouri et al.^47^, (2019) reported that the mean BLC was higher in young smokers than to older people in Birjand city, but this difference was no significant. The results of this study is inconsistent with our study, which could be either due to different population size or related to a different diet.

The previous studies have shown that while working with lead, frequent and proper use of appropriate PPE results in lower BLCs among workers ^48,49^. in the present study, The BLCs were higher among the welders who did not use appropriate PPE. This suggests that using appropriate PPE could be effective in reducing lead exposure.

Our study is one of the few studies carried out in Eastern Iran to determine the levels of lead among workers. The results of this study showed that the ALC could be a determining predictor of workers exposure to lead. To measure other sources of welders’ lead exposure, it suggested that other studies be performed; therefore conducting future studies on long-term exposure to high levels of lead, and with a larger population size is recommended.

## Conclusion

The results of the present study showed that the mean BLC of automobile welders in Birjand and the mean ALC corresponded to the standard of ACGIH; besides, the mean BLC of those who did not use PPE was higher than the mean BLC of other welders, and it was also higher in individuals over the age of 30 than those under this age. Therefore, to reduce lead exposure, the results can provide the information necessary for occupational health measures.

## Data Availability

All data produced in the present study are available upon reasonable request to the authors

## Authors Contributions

OM designed the study and also supervised the manuscript. AGH revised the manuscript. GHSH analyzed the data and performed statistical analysis too. FJ collaborated in data acquisition and manuscript preparation. HL performed literature search and prepared the manuscript; besides, all authors read and approved the final version of the manuscript.

## Acknowledgement

We sincerely thank the help of the Vice-Chancellor for Research for the study financing.

## Ethical Approval and Consent to Participate

This study was approved by the Committee on Research Ethics at Birjand University of Medical Sciences with the approved No. 4794 (IR.Bums.REC.1396.360).

## Data Availability

The datasets generated and analyzed during the current study are available from the corresponding author on reasonable request.

## Supplementary Data

The supplementary data are available on the journal’s website and the published article.

